# Debriefing to improve interprofessional teamwork in the operating room: a systematic review

**DOI:** 10.1101/2022.07.02.22277174

**Authors:** Emma Skegg, Canice McElroy, Mercedes Mudgway, James Hamill

## Abstract

**Purpose:** Debriefing has been pivotal in medical simulation training but its application to the real-world operating room environment has been challenging. We reviewed the literature on routine surgical debriefing with special reference to its implementation, barriers, and effectiveness.

**Design:** Descriptive systematic review following the Preferred Reporting Items for Systematic Reviews and Meta-Analyses guidelines.

**Methods:** Inclusion criteria were papers pertaining to debriefing in routine surgical practice. Excluded were papers reporting simulation training. We searched Google Scholar, CINAHL, Web of Science Core Collection, PsychINFO, Medline, Embase, and ProQuest Theses & Dissertations Global. The last search was performed on 14 March 2022. Quality was assessed on a 21-point checklist adapted from a standard reporting guideline. Synthesis was descriptive.

**Findings:** The search process resulted in 19 papers. Publication date ranged from 2007 – 2022. Study methods included surveys, interviews, and analysis of administrative data. Five papers involved a specific intervention. Quality scores ranged from 12 – 19 out of 21. On synthesis, we identified five topics: explanations of how debriefing had been implemented; the value of coaching and audit; the learning dimensions of debriefing, both team learning and quality improvement at the organizational level; the effect of debriefing on patient safety or the organization’s culture; and barriers to debriefing.

**Conclusions:** Debriefing is valuable for team learning, efficiency, patient safety, and psychological safety. Successful implementation programs were characterized by strong commitment from management and support by frontline workers. Integration with administrative quality and safety processes, and information feedback to frontline workers are fundamental to successful debriefing programs.

## Introduction

Errors in medicine have been recognized as a problem for over half a century. Donchin et al. (1995) found that, in their intensive care unit, only 0.95% of activities lead to an error but with 179 activities per patient per day there were on average 1.75 errors per patient per day (Donchin et al., 1995). “Active” errors (the effects of which are felt straight away) can almost always be linked to a series of “latent” errors (“accidents waiting to happen”) (Leape, 1994). Latent errors occur even during successful operations. Catchpole et al. (2006) showed that threats and errors in the operating room can come from organizational culture, processes, protocols, techniques, equipment, knowledge, skill or expertise, resources, anatomical variations or physiological problems with the patient, and problems with teamwork (Catchpole et al., 2006).

An important advance in surgical teamwork came with the WHO Surgical Safety Checklist (SSC) (Gawande, 2010; Martin et al., 2009). The SSC is a communication tool used at time points in the surgical process. These time points are the “sign-in” before anesthetizing, the “time-out” prior to the incision, and the “sign-out” at the end of the case. Hayes et al. (2009) showed that the SSC significantly reduces morbidity and mortality in a variety of high, middle and low-income settings (Haynes et al., 2009). Other components of the SSC that were not formally investigated in the study published by Hayes et al. (2009) were the briefing, which occurs before the operating list begins, and the debriefing at the end of a list.

Debriefing is a way to identify errors, improve performance, improve communication and promote teamwork (Zuckerman et al., 2012). The synonymous use of the terms “debriefing” and “sign-out” could cause confusion. In the present review, we emphasize the value of a comprehensive team discussion, that is, debriefing, over and above the previously established value of the sign-out checklist (Haynes et al., 2009). Operating room teams can be described as action teams (Vashdi et al., 2013). Action teams undertake time-critical, high-stakes active procedures. Operating room teams comprise personnel from a variety of professions and are therefore multidisciplinary action teams. High-level functioning of multidisciplinary action teams is critical and challenging. From a review of 20 years’ literature, Salas et al. (2005) identified the key characteristics of an effective team: leadership, mutual performance monitoring, backup behavior, adaptability, team orientation, shared mental models, mutual trust, and closed-loop communication (Salas et al., 2005). To achieve these, team members must feel safe about sharing observations and opinions with the rest of the team (Edmondson, 1999). Research shows that debriefing improves psychological safety in the operating room (Leong et al., 2017) and that psychological safety not only facilitates team learning, it helps maintain mental health and prevents burnout as well (Swendiman et al., 2019).

Given the central role that debriefing plays in teamwork, the experience in other industries and in medical simulation, and the importance of psychological safety, debriefing should be performed routinely in surgery. However, in our experience, debriefing is challenging to do well and is inconsistently performed. We were interested in learning how to best instigate a debriefing program. Therefore, the aim of this review was to synthesize the literature on routine surgical debriefing with specific reference to implementation, barriers, and the effectiveness of surgical debriefing, and to identify gaps in the literature that could indicate future research directions.

## Methods

This review and its protocol are registered on the Open Science Framework (https://osf.io/r5zba/).

### Eligibility Criteria

Criteria for including a paper in this review were studies pertaining to debriefing in routine practice. Excluded were papers pertaining to medical simulation training. No study design or language limits were imposed. No date limit was applied.

### Information Sources

We searched the databases Google Scholar, CINAHL, Web of Science Core Collection, PsychINFO, Medline, Embase, and ProQuest Theses & Dissertations Global. We performed snowballing and citation tracking by scanning the reference lists of included papers.

### Search Strategy

We used the following search teams in the database searches: debrief*, operating room*, operating theat*, surgical procedures and/or operative and/or operating rooms. For some searches, we added a title/abstract term: simulat*. The full search strategy for Medline is shown in Table 1. To be indexed, papers needed to mention at least one term related to each key variable: debrief and operating. The last date of searching was 14 March 2022.

**Table 1.**
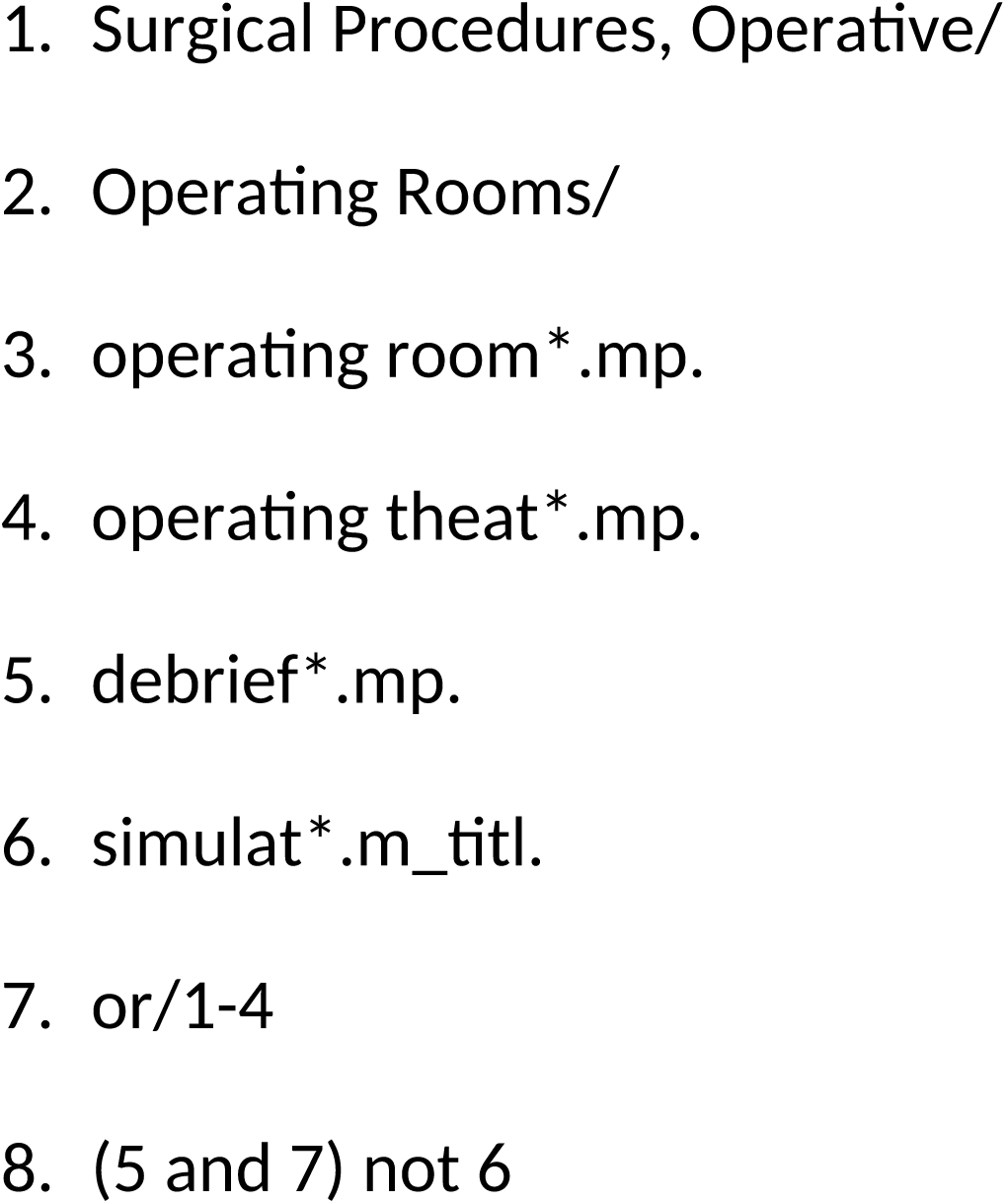
Search strategy for Medline

### Study Records

Literature search results were exported from each electronic database then imported into Rayyan (Ouzzani et al., 2016). MM removed duplicates. To ensure inter-rater reliability, two reviewers (MM and ES) independently screened titles and abstracts and removed any papers clearly not meeting inclusion criteria. Blinding was then turned off to deal with discrepancies which were resolved at a meeting with a third reviewer (JH). When the list of potentially included results was agreed, reviewers obtained the full-text reports. The same two reviewers independently reviewed the full texts for inclusion and resolved discrepancies at a meeting involving the same third reviewer.

### Data Items

Data were extracted on methods, how debriefing was implemented, the description of when and how debriefing was performed, any interventions, and outcomes. JH extracted data and MM checked data for accuracy.

### Synthesis

For descriptive analysis we grouped papers into five broad categories: 1) explanations of how debriefing had been implemented including how coaching had helped to improve the quantity and quality of debriefing; 2) the role of coaching and audit; 3) the learning dimensions of debriefing, both team learning and quality improvement at the organizational level; 4) the effect of debriefing on patient safety or the organization’s culture; and 5) the barriers to debriefing.

### Quality Assessment Methods

Quality assessment was by use of a customized checklist adapted from the Standards for Reporting Qualitative Research guidelines (O’Brien et al., 2014). An adapted checklist was used due to the variance of methods of included studies and although some included quantitative as well as qualitative data, outcomes were so variable that the application of standard quality assessment tools for systematic reviews was not feasible. Papers were scored on 21 items including the quality of their title, abstract, problem formulation, purpose, research paradigm, reflexivity of the researchers, context/setting, sampling, ethics, data collection methods, data collection instruments, units of study, data processing, data analysis, techniques to enhance trustworthiness, interpretation, links to empirical data, integration with prior work, discussion of limitations, and declarations of conflict of interest and funding. Items were scored as 1 or 0 for adequate or inadequate respectively.

## Results

### Description of Studies

The search process resulted in 19 papers as shown in the PRISMA flow diagram, Fig. 1. The characteristics of each included paper are presented in Table 2. Publication dates ranged from 2003 – 2022 with the majority (15 of 19) published in the last 10 years.

**Figure 1.**
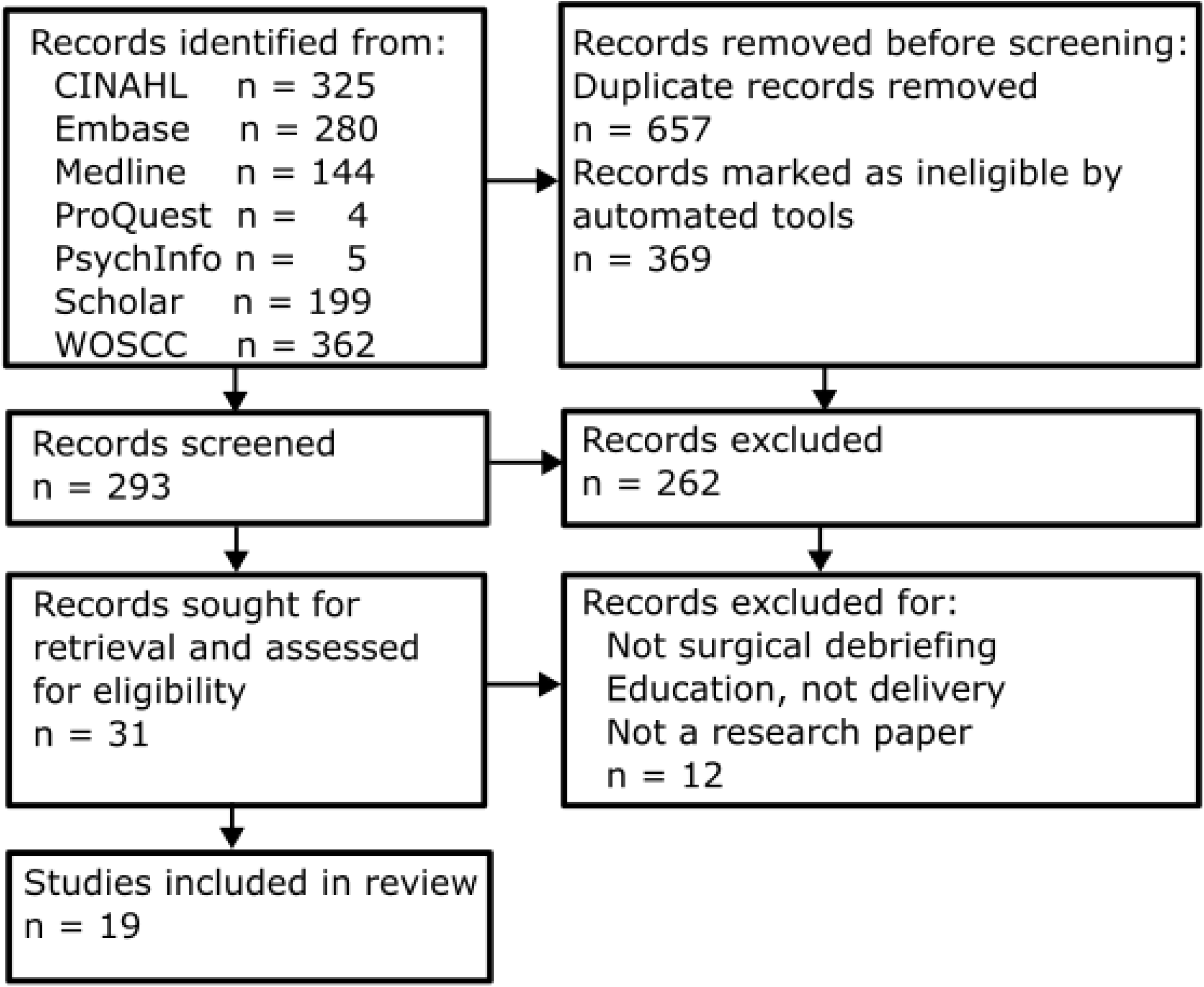
Prisma flow diagram.

**Table 2.**
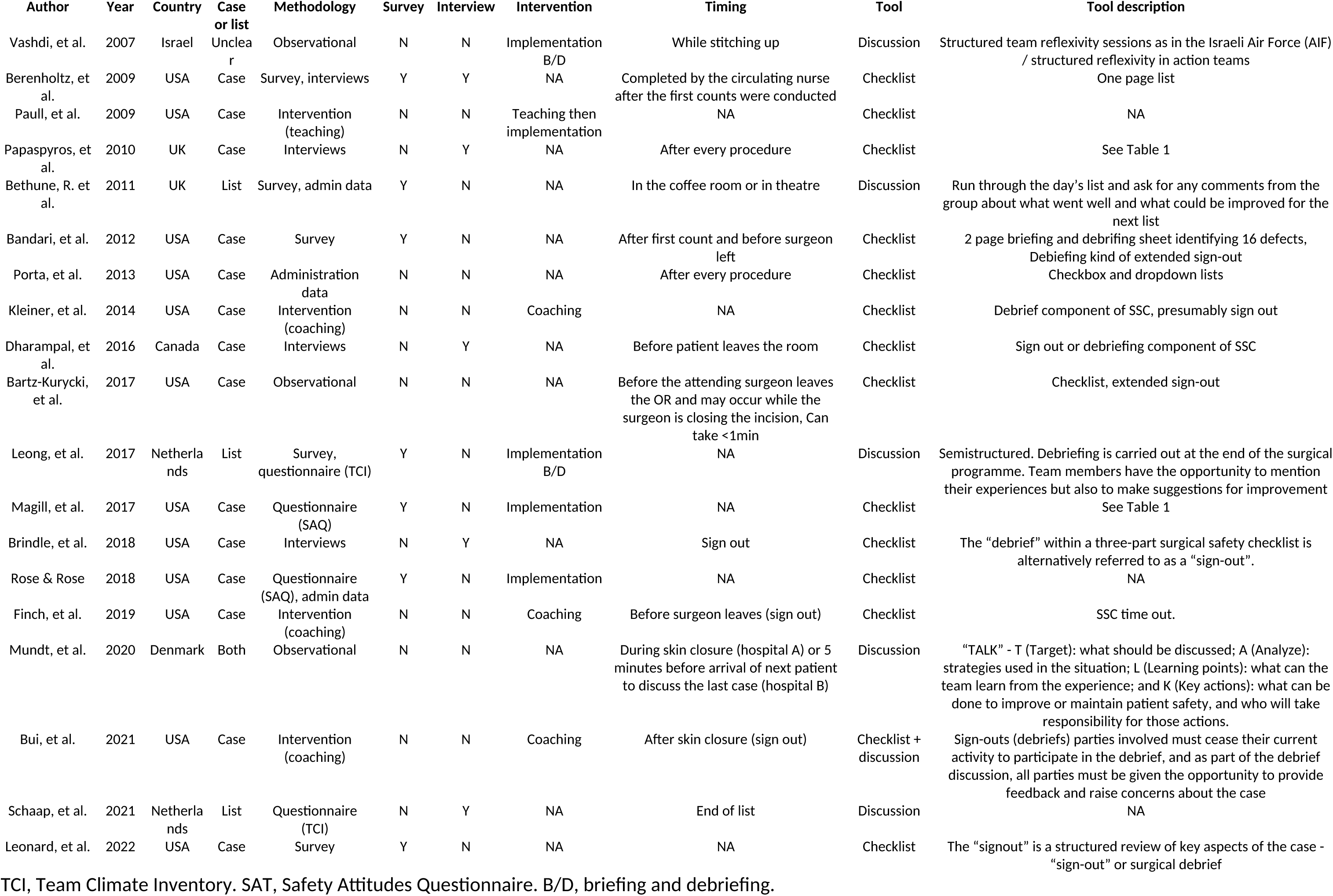
Papers included in the review with baseline data

Most papers (n = 15) referred to debriefing as a form of “sign-out” checklist rather than a dedicated team discussion at the end of an operating session. One study (Mundt et al., 2020) involved the orthopedic departments of two hospitals in which one unit debriefed after every case and the other unit debriefed at the end of the operating list. Seven papers involved an intervention: implementation of a debriefing (and briefing) process in five (Leong et al., 2017; Magill et al., 2017; Paull et al., 2009; Rose & Rose, 2018; Vashdi et al., 2013) and a coaching intervention to improve the quality of debriefs in three (Bui et al., 2021; Finch et al., 2019; Kleiner et al., 2014).

### Assessment of Quality

The mean quality score was 15/21 (standard deviation 2.1, range 12 – 19). All papers met criteria for title (100%), abstract (100%), problem formulation (100%), purpose or research question (100%), context (100%), data collection methods (100%), data collection instruments and technologies (100%), synthesis and interpretation (100%), and integration with prior work, implications, transferability, and contribution(s) to the field (100%). Few reported the qualitative approach or research paradigm (37%), ethical issues pertaining to human subjects (47%), or the process by which themes were developed (32%). No paper reported researcher characteristics/reflexivity or techniques to enhance trustworthiness. Quality assessments for each paper are presented in Table S1 in Supporting Information.

### 1. Implementation of Debriefing

Seven papers reported on the implementation of debriefing in five different hospitals (Bandari et al., 2012; Berenholtz et al., 2009; Bethune et al., 2011; Brindle et al., 2018; Mundt et al., 2020; Papaspyros et al., 2010; Rose & Rose, 2018). Implementation programs were of two types: top-down, where hospital clinicians and administration worked together to bring in a briefing/debriefing practice; and grassroots, where clinicians and researchers developed a debriefing practice in their workplace. Top-down implementation programs were characterized by management taking active leadership in developing meaningful links to quality improvement processes and involving surgical and nursing staff as described by Rose and Rose (2018) and further explored by Brindle et al. (2018). Rose and Rose (2018) describe this as a “multilevel, multipronged” approach.

An example of successful top-down implementation was McLeod Regional Medical Center, where the team involved “stakeholders at every organizational level in crafting and implementing solutions” (Rose & Rose, 2018), fed back all their findings to staff, executive leadership and the governing board, and “set accountability for management to analyze events, follow up on findings, and drive change” (Rose & Rose, 2018). They attribute their success to executive staff being physically present in the operating room, a culture of safety, a just culture that focused on empowering nurses and encouraging open communication, leveling the playing field, and providing caregivers with early and meaningful feedback on the issues they had raised, the latter being regarded as the cornerstone to successful debriefing (Brindle et al., 2018). An example of top-down implementation that eventually failed was Beaumont Hospital, Michigan. The hospital implemented a multidisciplinary team of frontline theatre staff, administrators, and hospital leadership, to launch a briefing/debriefing program across a range of surgical specialties (Bandari et al., 2012; Berenholtz et al., 2009). The authors emphasized the importance of buy-in from clinicians:

“When implementing the briefing and debriefing tool, we learned a few important lessons. Gaining physician and staff buy-in was the single most significant barrier.” − (Berenholtz et al., 2009)

After a change in executive leadership, the system collapsed. The hospital leadership changed from a hands-on to an arms-length approach, and removed the quality and safety nurse who had been dealing with issues raised during debriefings and had providing feedback to caregivers. Loss of feedback was seen as the key aspect that led to the program’s demise (Brindle et al., 2018).

Grassroots implementation programs were characterized by clinician and researcher leadership. Bethune et al. (2011) described a less successful experience of grassroots implementation. The researchers encouraged staff to gather after operating lists to debrief; however, the process was found to be difficult to implement due to the lack of buy-in from senior staff. There is no mention of a multi-disciplinary consultation process that could have have aided this issue (Bethune et al., 2011). In another paper, Mundt et al. (2020) reported setting up debriefings led by trained facilitators. The authors mentioned close collaboration with heads of department but no system to link debrief outcomes with quality improvement. Participants cited problems with the feasibility of performing debriefs (Mundt et al., 2020). In a third paper, Papaspyros et al. (2010) reported recurring issues coming up in debriefs that were not resolved despite following standard hospital reporting procedures. Successful implementation involved developing a culture of safety, leveling the playing field, empowering nurses, open communication, and providing caregivers with early and meaningful feedback on the issues they had raised. Unsuccessful implementation was characterized by lack of managerial leadership, an “arms-length” approach, not dealing with issues raised, and loss of communication and feedback to frontline staff (Brindle et al., 2018).

Debriefing initiatives that were not characterized by multidisciplinary, multilevel leadership suffered from lack of buy-in from senior clinicians (Bethune et al., 2011), no link to quality improvement, and lack of communication and feedback to debriefers (Mundt et al., 2020; Papaspyros et al., 2010). Together, these papers show that successful implementation requires debriefing to be resourced, integrated at all levels from grassroots to executive, and integrated with quality processes.

### 2. Coaching and Audit

Three papers described how debriefing improved with coaching (Bui et al., 2021; Finch et al., 2019; Kleiner et al., 2014). In one study (Kleiner et al., 2014), a retired surgeon debriefed staff on the quality of their debriefs, as well as conducting group discussions and presentations. The researchers found a significant improvement in the quality of debriefs after coaching (Kleiner et al., 2014). In a second study (Finch et al., 2019) two coaches met with nurses and surgeons, either as a group or individually. The coaches, nurse managers and educators, and anesthetists audited debriefs before and after the coaching intervention using standardized audit tools. The researchers found an increase in the number of debriefs performed and the completeness of debriefs after coaching. These authors emphasized auditing as an important part of their implementation project (Finch et al., 2019). In a third study (Bui et al., 2021), the researchers gave surgeons structured feedback on the quality of their debriefs and provided one-on-one coaching for surgeons who were finding debriefing difficult. The number of debriefs performed and the completeness of debriefs increased after coaching (Bui et al., 2021). Two further papers (Paull et al., 2009; Vashdi et al., 2013) looked at external education or guidance in debriefing. Paull et al. (2009) ran teaching sessions for leaders who would implement a new briefing/debriefing practice in their own hospitals. The authors found that participants who engaged the most with the teaching process also had the most success with implementation. Vashdi et al. (2013) engaged with debriefing experts from the Air Force to bring military briefing/debriefing methods into the surgical units of a teaching hospital. Coaching and feedback increased the number of debriefs performed and their quality and completeness. Finch et al. (2019) emphasized auditing debriefs as an important way to maintain compliance and completeness. Learning from others in the form of an education course (Paull et al., 2009) and using the experience of an outside organization (Vashdi et al., 2007) were also seen as valuable. Together, these papers show that debriefing benefits from guidance and that debriefing programs should include ongoing coaching and support.

### 3. Team and Institutional Learning

Investigators for one study took an in-depth look at team learning processes as they relate to debriefing (Vashdi et al., 2013). The authors applied the brief/debrief practices of the Israeli Air Force to surgical units in a tertiary hospital, with special focus on team learning. Surgery was seen to have problems with an unhealthy approach to criticism, status-based relationships between professional groups that impaired communication, difficulties in scheduling meetings, and inhibitions arising from concerns about liability. The research team proposed mechanisms by which surgical teams could improve from one surgery to the next. These were: single-loop learning (“problems identified in the context of a debriefing for a surgery conducted at time T1 are translated into improvements in planned actions included in the preoperative briefing for surgeries to be conducted at time T2” (Vashdi et al., 2013)); direct double-loop learning (“engage in the questioning of taken-for-granted procedures and the assumptions underlying them’ and “search for the common, system-level roots of recurring problems” (Vashdi et al., 2013)); and indirect double-loop learning (“the effect of structured team reflexivity on quality-related surgical team outcomes will be mediated by the degree to which surgical team members share a common sense of the importance of inquiry, transparency, and accountability in their day-to-day team-based interactions” (Vashdi et al., 2013)).

Several papers addressed the potential for debriefing to facilitate institutional learning through quality improvement processes (Bandari et al., 2012; Brindle et al., 2018; Rose & Rose, 2018; Vashdi et al., 2007). In one study (Rose & Rose, 2018) clinical, finance and supply chain managers reviewed all problems identified by debriefs to determine root causes, recommend solutions, and allocate actions, analysis and review to management team members. More than half of the problems identified in debriefs were of an institutional nature − problems with workflow, room readiness, instruments or the supply chain − indicating that surgical teams would be powerless to influence most of the issues identified in their debriefs if management were not active participants in the process. As the authors put it, “large-scale improvement only became possible with the tight coordination and contribution of scores of people around the surgery event” (Rose & Rose, 2018). Another study (Vashdi et al., 2013) described weekly cross-team departmental meetings to review the previous week’s debriefings, looking for patterns that might indicate the need for system-level change. Together, these papers emphasize the importance of communicating back to the team the actions that had been undertaken as a result of their debriefs.

### 4. The Effects of Debriefing on Culture, Safety, and Efficiency

Several papers reported on the effects of debriefing on culture, quality and safety. Rose and Rose (2018) showed a significant reduction in postoperative mortality after the implementation of a debriefing program. The authors emphasized the need for long-term thinking because improvements occur over a period of years. In two papers (Porta et al., 2013; Rose & Rose, 2018), debriefing improved efficiency as measured by a reduction in delay to the operating room, increased utilization, more accurate scheduling of operations (Porta et al., 2013), and reduced staff working hours per case (Rose & Rose, 2018). Debriefing was shown to improve the climate of psychological safety in several papers. Leonard et al. (2022) found that debriefing helped operating room workers to feel more comfortable when speaking up; this applied particularly to circulating nurses, scrub technicians, and anesthetic advanced practice providers.

Two papers (Magill et al., 2017; Rose & Rose, 2018) showed that scores on the Safety Attitudes Questionnaire (Sexton et al., 2006) improved with debriefing. Similarly, two papers (Leong et al., 2017; Schaap et al., 2021) showed significantly increased scores on the Team Climate Inventory (Anderson & West, 1998) after a briefing/debriefing program was introduced. Together, these papers show that debriefing improved safety for patients and staff.

### 5. Barriers to Debriefing

Power dynamics may raise barriers to debriefing (Leonard et al., 2022). In a survey of operating room staff, most nurses and technicians thought that the responsibility for creating a culture of safety was a shared responsibility; however, over one half of surgeons thought it was the surgeon’s responsibility (Leonard et al., 2022). Nurses and technicians felt more strongly that debriefing helped to create a culture of speaking up compared to surgeons (Leonard et al., 2022). Time presented a challenge to debriefing (Bethune et al., 2011; Mundt et al., 2020; Schaap et al., 2021). Operating room staff reported timing difficulties with debriefing at the end of short cases, preferring to debrief at the end of the whole list (Mundt et al., 2020). Debriefing near the end of a procedure can present difficulties if surgeons need to concentrate on operating or anesthetists need to focus on reversal of anesthesia (Dharampal et al., 2016). Debriefing between cases presented difficulties with getting people together again after the case (Bethune et al., 2011; Mundt et al., 2020; Schaap et al., 2021). Sometimes, senior surgeons would have left the operation before the debrief, allowing more junior staff to finish (Dharampal et al., 2016). A further barrier to debriefing was management’s inability or unwillingness to make changes that would enable routine debriefing (Rose & Rose, 2018). Repeated, unresolved problems and thwarted efforts to improve processes undermine the credibility of the process and can be exhausting for staff (Rose & Rose, 2018). This shows the need for “top-down” leadership by managers to complement enthusiasm at the grassroots level. Lack of buy-in, perceiving little benefit, thinking everything went well so there is nothing to debrief on, and not seeing debriefing as a priority were other barriers identified (Schaap et al., 2021). Together, these papers show that team orientation and support at all levels facilitates debriefing while hierarchy, time pressure, lack of buy-in and inaction on issues represent barriers.

## Discussion

This review shows that the literature on surgical debriefing is relatively sparse and not generally of high quality; nevertheless, the literature offers guidance on how to implement a debriefing program, the value of coaching in implementation and maintenance, insights into team learning, lessons on the need to take a systems-wide view of quality and improvement, and data on the effect of debriefing on teams and on patient outcomes. Collaboration between clinicians, management and quality services is a prerequisite for a successful debriefing program. The most successful programs had strong leadership from the hospital administration, good governance and took time and commitment. It was interesting to contrast the examples of successful implementation with the unsuccessful in which evidence of collaboration was lacking. The success of coaching in supporting debriefing programs was further evidence of the need for commitment by leadership.

The translation of a military briefing/debriefing practice into an operating room setting provided interesting insights into how much potential there is for team learning to improve in surgery, but also the challenges in the medical setting. Many of these challenges are deeply cultural in nature, such as the culture of hierarchy. Addressing barriers in order to bring about debriefing and through the *use* of debriefing would enhance patient care as well as work satisfaction.

Teams that learn well perform well. This is especially important in surgery where teams must adapt quickly in critical situations. The operating room environment provides a rich source of experience, but experience alone is not enough for effective learning (Mayer, 2004). Learning involves a cycle of concrete experience, reflective observation, abstract hypotheses, and active testing (Kolb & Kolb, 2006). Learning occurs within learning spaces, not necessarily just physical spaces but also constructs in the social environment (Kolb & Kolb, 2006). Given the potential for operating room teams to continually learn and improve it is surprising that team learning processes such as debriefing are not routine.

Briefing and debriefing were often addressed together in the papers in this review. Briefing is a team meeting that occurs at the beginning of an operation or operating list, while debriefing is a team discussion that occurs after the operation or operating list. From a learning perspective, both discussions go hand-in-hand; however, debriefing appears to be more difficult to enact than briefing. Barriers to debriefing included time pressures, not being able to get the whole team together after a case, and difficulties with buy-in from some staff. The timing problem may explain why some papers in this review located the debrief at the sign-out (Bartz-Kurycki et al., 2017; Dharampal et al., 2016; Magill et al., 2017). The sign-out is the third of the Surgical Safety Checklists and occurs during the completion of a case, usually after the count (which ensures all instruments and swabs are accounted for).

## Limitations

This review has several limitations. We included papers that reported debriefing but, on review, were using the term “debrief” as a synonym for the sign-out phase of the Surgical Safety Checklist; thus, we may have overestimated the number of relevant papers available. Many papers reported debriefing on a case-basis (i.e., after every operative case) and they were clearly on the spectrum as to how much of the debrief was a team discussion versus a checklist. Meta-analysis was not possible given the lack of data and variability of outcomes, limiting this to a descriptive review.

## Conclusions

In conclusion, debriefing appears to be valuable for team learning, efficiency, patient safety and psychological safety. Surgical debriefing is challenging to implement and maintain. Successful programs are characterized by strong commitment from management in addition to support by frontline workers. Integration with administrative quality and safety processes and feedback to frontline workers are fundamental to a successful debriefing program. Overall, literature is lacking on surgical debriefing and more research on implementation, maintenance, and outcomes are required.

## Data Availability

All data produced in the present work are contained in the manuscript and supplementary material

## Notes

Declarations Emma Skegg and Mercedes Mudgway were funded by a grant from the Starship Foundation (SF2142). The authors have no conflicts of interest to declare.

### Competing Interest Statement

The authors have declared no competing interest.

### Clinical Protocols

https://osf.io/r5zba/

### Funding Statement

Emma Skegg and Mercedes Mudgway were funded by a grant from the Starship Foundation (SF2142).

### Summary of Updates

Revisions of some of the text for clarity and to remove duplicate statements.

