## Supplementary material for "Debriefing to improve interprofessional teamwork in the operating room: a systematic review": Table S1

Table S1. Quality scores for papers included in the review

| **Article Name and Author: Study 1** | **The surgical debrief: Just another checklist or an instrument to drive cultural change?** | **Leonard et al. (2021)** |
|---|---|---|
| **Title** – Concise description of the nature and topic of the study Identifying the study as qualitative or indicating the approach (e.g., ethnography, grounded theory) or data collection methods (e.g., interview, focus group) is recommended | Yes | |
| **Abstract** - Summary of key elements of the study using the abstract format of the intended publication; typically includes background, purpose, methods, results, and conclusions | Yes | |
| **Problem formulation** - Description and significance of the problem/phenomenon studied; review of relevant theory and empirical work; problem statement | Yes | |
| **Purpose or research questio**n - Purpose of the study and specific objectives or questions | Yes | |
| **Qualitative approach and research paradigm** - Qualitative approach (e.g., ethnography, grounded theory, case study, phenomenology, narrative research) and guiding theory if appropriate; identifying the research paradigm (e.g., postpositivist, constructivist/ interpretivist) is also recommended; rationale** | No | |
| **Researcher characteristics and reflexivity** - Researchers' characteristics that may influence the research, including personal attributes, qualifications/experience, relationship with participants, assumptions, and/or presuppositions; potential or actual interaction between researchers' characteristics and the research questions, approach, methods, results, and/or transferability | No | |
| **Context** - Setting/site and salient contextual factors; rationale** | Yes | |
| **Sampling strategy** - How and why research participants, documents, or events were selected; criteria for deciding when no further sampling was necessary (e.g., sampling saturation); rationale** | Yes | |
| **Ethical issues pertaining to human subjects** - Documentation of approval by an appropriate ethics review board and participant consent, or explanation for lack thereof; other confidentiality and data | No | |

| | |
|---|---|
| security issues | |
| **Data collection methods -** Types of data collected; details of data collection procedures including (as appropriate) start and stop dates of data collection and analysis, iterative process, triangulation of sources/methods, and modification of procedures in response to evolving study findings; rationale** | Yes |
| **Data collection instruments and technologies -** Description of instruments (e.g., interview guides, questionnaires) and devices (e.g., audio recorders) used for data collection; if/how the instrument(s) changed over the course of the study | Yes |
| **Units of study -** Number and relevant characteristics of participants, documents, or events included in the study; level of participation (could be reported in results) | Yes |
| **Data processing -** Methods for processing data prior to and during analysis, including transcription, data entry, data management and security, verification of data integrity, data coding, and anonymization/de-identification of excerpts | Yes |
| **Data analysis -** Process by which inferences, themes, etc., were identified and developed, including the researchers involved in data analysis; usually references a specific paradigm or approach; rationale** | No |
| **Techniques to enhance trustworthiness -** Techniques to enhance trustworthiness and credibility of data analysis (e.g., member checking, audit trail, triangulation); rationale** | No |
| **Synthesis and interpretation -** Main findings (e.g., interpretations, inferences, and themes); might include development of a theory or model, or integration with prior research or theory | Yes |
| **Links to empirical data -** Evidence (e.g., quotes, field notes, text excerpts, photographs) to substantiate analytic findings | Yes |
| **Integration with prior work, implications, transferability, and contribution(s) to the field -** Short summary of main findings; explanation of how findings and conclusions connect to, support, elaborate on, or challenge conclusions of earlier scholarship; discussion of scope of application/generalizability; identification of unique contribution(s) to scholarship in a discipline or field | Yes |

| | | |
|---|---|---|
| **Limitations** - Trustworthiness and limitations of findings | Yes | |
| **Conflicts of interest** - Potential sources of influence or perceived influence on study conduct and conclusions; how these were managed | No | |
| **Funding** - Sources of funding and other support; role of funders in data collection, interpretation, and reporting | Yes | 15/21 |
| **Article Name and Author: Study 2** | **Structured feedback enhances compliance with operating room debriefs** | **But et al. (2020)** |
| **Title** - Concise description of the nature and topic of the study Identifying the study as qualitative or indicating the approach (e.g., ethnography, grounded theory) or data collection methods (e.g., interview, focus group) is recommended | Yes | |
| **Abstract** - Summary of key elements of the study using the abstract format of the intended publication; typically includes background, purpose, methods, results, and conclusions | Yes |  |
| **Problem formulation** - Description and significance of the problem/phenomenon studied; review of relevant theory and empirical work; problem statement | Yes | |
| **Purpose or research questio**n - Purpose of the study and specific objectives or questions | Yes | |
| **Qualitative approach and research paradigm** - Qualitative approach (e.g., ethnography, grounded theory, case study, phenomenology, narrative research) and guiding theory if appropriate; identifying the research paradigm (e.g., postpositivist, constructivist/ interpretivist) is also recommended; rationale** | No | |
| **Researcher characteristics and reflexivity** - Researchers' characteristics that may influence the research, including personal attributes, qualifications/experience, relationship with participants, assumptions, and/or presuppositions; potential or actual interaction between researchers' characteristics and the research questions, approach, methods, results, and/or transferability | No | |
| **Context** - Setting/site and salient contextual factors; rationale** | Yes | |
| **Sampling strategy** - How and why research participants, documents, or events were selected; | Yes | |

| | |
|---|---|
| criteria for deciding when no further sampling was necessary (e.g., sampling saturation); rationale** | |
| **Ethical issues pertaining to human subjects -** Documentation of approval by an appropriate ethics review board and participant consent, or explanation for lack thereof; other Yesconfidentiality and data security issues | No |
| **Data collection methods -** Types of data collected; details of data collection procedures including (as appropriate) start and stop dates of data collection and analysis, iterative process, triangulation of sources/methods, and modification of procedures in response to evolving study findings; rationale** | Yes |
| **Data collection instruments and technologies -** Description of instruments (e.g., interview guides, questionnaires) and devices (e.g., audio recorders) used for data collection; if/how the instrument(s) changed over the course of the study | Yes |
| **Units of study -** Number and relevant characteristics of participants, documents, or events included in the study; level of participation (could be reported in results) | Yes |
| **Data processing -** Methods for processing data prior to and during analysis, including transcription, data entry, data management and security, verification of data integrity, data coding, and anonymization/de-identification of excerpts | Yes |
| **Data analysis -** Process by which inferences, themes, etc., were identified and developed, including the researchers involved in data analysis; usually references a specific paradigm or approach; rationale** | No |
| **Techniques to enhance trustworthiness -** Techniques to enhance trustworthiness and credibility of data analysis (e.g., member checking, audit trail, triangulation); rationale** | No |
| **Synthesis and interpretation -** Main findings (e.g., interpretations, inferences, and themes); might include development of a theory or model, or integration with prior research or theory | Yes |
| **Links to empirical data -** Evidence (e.g., quotes, field notes, text excerpts, photographs) to substantiate analytic findings | Yes |
| **Integration with prior work, implications, transferability, and contribution(s) to the field -** | Yes |

| | | |
|---|---|---|
| Short summary of main findings; explanation of how findings and conclusions connect to, support, elaborate on, or challenge conclusions of earlier scholarship; discussion of scope of application/generalizability; identification of unique contribution(s) to scholarship in a discipline or field | | |
| **Limitations** - Trustworthiness and limitations of findings | Yes | |
| **Conflicts of interest -** Potential sources of influence or perceived influence on study conduct and conclusions; how these were managed | Yes | |
| **Funding** - Sources of funding and other support; role of funders in data collection, interpretation, and reporting | Yes | 16/21 |

| **Article Name and Author: Study 3** | **Debriefing: the forgotten phase of the surgical safety checklist** | **Bartz-Kurycki et al. (2017)** |
|---|---|---|
| **Title -** Concise description of the nature and topic of the study Identifying the study as qualitative or indicating the approach (e.g., ethnography, grounded theory) or data collection methods (e.g., interview, focus group) is recommended | Yes | |
| **Abstract** - Summary of key elements of the study using the abstract format of the intended publication; typically includes background, purpose, methods, results, and conclusions | Yes | |
| **Problem formulation** - Description and significance of the problem/phenomenon studied; review of relevant theory and empirical work; problem statement | Yes | |
| **Purpose or research questio**n - Purpose of the study and specific objectives or questions | Yes | |
| **Qualitative approach and research paradigm** - Qualitative approach (e.g., ethnography, grounded theory, case study, phenomenology, narrative research) and guiding theory if appropriate; identifying the research paradigm (e.g., postpositivist, constructivist/ interpretivist) is also recommended; rationale** | No | |
| **Researcher characteristics and reflexivity** - Researchers' characteristics that may influence the research, including personal attributes, qualifications/experience, relationship with participants, assumptions, and/or presuppositions; potential or actual interaction between researchers' characteristics and the research questions, | No | |

| | |
|---|---|
| approach, methods, results, and/or transferability | |
| **Context** - Setting/site and salient contextual factors; rationale** | Yes |
| **Sampling strategy** - How and why research participants, documents, or events were selected; criteria for deciding when no further sampling was necessary (e.g., sampling saturation); rationale** | Yes |
| **Ethical issues pertaining to human subjects** - Documentation of approval by an appropriate ethics review board and participant consent, or explanation for lack thereof; other confidentiality and data security issues | No |
| **Data collection methods** - Types of data collected; details of data collection procedures including (as appropriate) start and stop dates of data collection and analysis, iterative process, triangulation of sources/methods, and modification of procedures in response to evolving study findings; rationale** | Yes |
| **Data collection instruments and technologies** - Description of instruments (e.g., interview guides, questionnaires) and devices (e.g., audio recorders) used for data collection; if/how the instrument(s) changed over the course of the study | Yes |
| **Units of study** - Number and relevant characteristics of participants, documents, or events included in the study; level of participation (could be reported in results) | Yes |
| **Data processing** - Methods for processing data prior to and during analysis, including transcription, data entry, data management and security, verification of data integrity, data coding, and anonymization/de-identification of excerpts | Yes |
| **Data analysis** - Process by which inferences, themes, etc., were identified and developed, including the researchers involved in data analysis; usually references a specific paradigm or approach; rationale** | No |
| **Techniques to enhance trustworthiness** - Techniques to enhance trustworthiness and credibility of data analysis (e.g., member checking, audit trail, triangulation); rationale** | No |
| **Synthesis and interpretation** - Main findings (e.g., interpretations, inferences, and themes); might include development of a theory or model, or integration with prior research or theory | Yes |

| | | |
|---|---|---|
| **Links to empirical data** - Evidence (e.g., quotes, field notes, text excerpts, photographs) to substantiate analytic findings | No | |
| **Integration with prior work, implications, transferability, and contribution(s) to the field -** Short summary of main findings; explanation of how findings and conclusions connect to, support, elaborate on, or challenge conclusions of earlier scholarship; discussion of scope of application/generalizability; identification of unique contribution(s) to scholarship in a discipline or field | Yes | |
| **Limitations** - Trustworthiness and limitations of findings | Yes | |
| **Conflicts of interest** - Potential sources of influence or perceived influence on study conduct and conclusions; how these were managed | No | |
| **Funding** - Sources of funding and other support; role of funders in data collection, interpretation, and reporting | No | 13/21 |

| **Article Name and Author: Study 4** | **Attitudes and beliefs about the surgical safety checklist: Just another tick box?** | **Dharampal et al. (2016)** |
|---|---|---|
| **Title** - Concise description of the nature and topic of the study Identifying the study as qualitative or indicating the approach (e.g., ethnography, grounded theory) or data collection methods (e.g., interview, focus group) is recommended | Yes | |
| **Abstract** - Summary of key elements of the study using the abstract format of the intended publication; typically includes background, purpose, methods, results, and conclusions | Yes | |
| **Problem formulation** - Description and significance of the problem/phenomenon studied; review of relevant theory and empirical work; problem statement | Yes | |
| **Purpose or research questio**n - Purpose of the study and specific objectives or questions | Yes | |
| **Qualitative approach and research paradigm** - Qualitative approach (e.g., ethnography, grounded theory, case study, phenomenology, narrative research) and guiding theory if appropriate; identifying the research paradigm (e.g., postpositivist, constructivist/ interpretivist) is also recommended; rationale** | Yes | |
| **Researcher characteristics and reflexivity -** | No | |

| | |
|---|---|
| Researchers’ characteristics that may influence the research, including personal attributes, qualifications/experience, relationship with participants, assumptions, and/or presuppositions; potential or actual interaction between researchers’ characteristics and the research questions, approach, methods, results, and/or transferability | |
| **Context** - Setting/site and salient contextual factors; rationale** | Yes |
| **Sampling strategy** - How and why research participants, documents, or events were selected; criteria for deciding when no further sampling was necessary (e.g., sampling saturation); rationale** | Yes |
| **Ethical issues pertaining to human subjects** - Documentation of approval by an appropriate ethics review board and participant consent, or explanation for lack thereof; other confidentiality and data security issues | Yes |
| **Data collection methods** - Types of data collected; details of data collection procedures including (as appropriate) start and stop dates of data collection and analysis, iterative process, triangulation of sources/methods, and modification of procedures in response to evolving study findings; rationale** | Yes |
| **Data collection instruments and technologies** - Description of instruments (e.g., interview guides, questionnaires) and devices (e.g., audio recorders) used for data collection; if/how the instrument(s) changed over the course of the study | Yes |
| **Units of study** - Number and relevant characteristics of participants, documents, or events included in the study; level of participation (could be reported in results) | Yes |
| **Data processing** - Methods for processing data prior to and during analysis, including transcription, data entry, data management and security, verification of data integrity, data coding, and anonymization/de-identification of excerpts | Yes |
| **Data analysis** - Process by which inferences, themes, etc., were identified and developed, including the researchers involved in data analysis; usually references a specific paradigm or approach; rationale** | Yes |
| **Techniques to enhance trustworthiness** - Techniques to enhance trustworthiness and | No |

| | | |
|---|---|---|
| credibility of data analysis (e.g., member checking, audit trail, triangulation); rationale** | | |
| **Synthesis and interpretation -** Main findings (e.g., interpretations, inferences, and themes); might include development of a theory or model, or integration with prior research or theory | Yes | |
| **Links to empirical data -** Evidence (e.g., quotes, field notes, text excerpts, photographs) to substantiate analytic findings | Yes | |
| **Integration with prior work, implications, transferability, and contribution(s) to the field -** Short summary of main findings; explanation of how findings and conclusions connect to, support, elaborate on, or challenge conclusions of earlier scholarship; discussion of scope of application/generalizability; identification of unique contribution(s) to scholarship in a discipline or field | Yes | |
| **Limitations -** Trustworthiness and limitations of findings | Yes | |
| **Conflicts of interest -** Potential sources of influence or perceived influence on study conduct and conclusions; how these were managed | Yes | |
| **Funding -** Sources of funding and other support; role of funders in data collection, interpretation, and reporting | Yes | 19/21 |

| **Article Name and Author: Study 5** | **Operating room efficiency improvement after implementation of a postoperative team assessment** | **Porta et al. (2013)** |
|---|---|---|
| **Title -** Concise description of the nature and topic of the study Identifying the study as qualitative or indicating the approach (e.g., ethnography, grounded theory) or data collection methods (e.g., interview, focus group) is recommended | Yes | |
| **Abstract -** Summary of key elements of the study using the abstract format of the intended publication; typically includes background, purpose, methods, results, and conclusions | Yes | |
| **Problem formulation -** Description and significance of the problem/phenomenon studied; review of relevant theory and empirical work; problem statement | Yes | |
| **Purpose or research question** - Purpose of the study and specific objectives or questions | Yes | |
| **Qualitative approach and research paradigm -** Qualitative approach (e.g., ethnography, grounded | No | |

| | |
|---|---|
| theory, case study, phenomenology, narrative research) and guiding theory if appropriate; identifying the research paradigm (e.g., postpositivist, constructivist/ interpretivist) is also recommended; rationale** | |
| **Researcher characteristics and reflexivity -** Researchers' characteristics that may influence the research, including personal attributes, qualifications/experience, relationship with participants, assumptions, and/or presuppositions; potential or actual interaction between researchers' characteristics and the research questions, approach, methods, results, and/or transferability | No |
| **Context -** Setting/site and salient contextual factors; rationale** | Yes |
| **Sampling strategy -** How and why research participants, documents, or events were selected; criteria for deciding when no further sampling was necessary (e.g., sampling saturation); rationale** | No |
| **Ethical issues pertaining to human subjects -** Documentation of approval by an appropriate ethics review board and participant consent, or explanation for lack thereof; other confidentiality and data security issues | Yes |
| **Data collection methods -** Types of data collected; details of data collection procedures including (as appropriate) start and stop dates of data collection and analysis, iterative process, triangulation of sources/methods, and modification of procedures in response to evolving study findings; rationale** | Yes |
| **Data collection instruments and technologies -** Description of instruments (e.g., interview guides, questionnaires) and devices (e.g., audio recorders) used for data collection; if/how the instrument(s) changed over the course of the study | Yes |
| **Units of study -** Number and relevant characteristics of participants, documents, or events included in the study; level of participation (could be reported in results) | Yes |
| **Data processing -** Methods for processing data prior to and during analysis, including transcription, data entry, data management and security, verification of data integrity, data coding, and anonymization/de-identification of excerpts | Yes |
| **Data analysis -** Process by which inferences, themes, | No |

| | | |
|---|---|---|
| etc., were identified and developed, including the researchers involved in data analysis; usually references a specific paradigm or approach; rationale** | | |
| **Techniques to enhance trustworthiness -** Techniques to enhance trustworthiness and credibility of data analysis (e.g., member checking, audit trail, triangulation); rationale** | No | |
| **Synthesis and interpretation -** Main findings (e.g., interpretations, inferences, and themes); might include development of a theory or model, or integration with prior research or theory | Yes | |
| **Links to empirical data -** Evidence (e.g., quotes, field notes, text excerpts, photographs) to substantiate analytic findings | Yes | |
| **Integration with prior work, implications, transferability, and contribution(s) to the field -** Short summary of main findings; explanation of how findings and conclusions connect to, support, elaborate on, or challenge conclusions of earlier scholarship; discussion of scope of application/generalizability; identification of unique contribution(s) to scholarship in a discipline or field | Yes | |
| **Limitations -** Trustworthiness and limitations of findings | No | |
| **Conflicts of interest -** Potential sources of influence or perceived influence on study conduct and conclusions; how these were managed | No | |
| **Funding -** Sources of funding and other support; role of funders in data collection, interpretation, and reporting | No | 13/21 |
| **Article Name and Author: Study 6** | **Briefing and debriefing in the cardiac operating room. Analysis of impact on theatre team attitude and patient safety** | **Papaspyros et al. (2009)** |
| **Title -** Concise description of the nature and topic of the study Identifying the study as qualitative or indicating the approach (e.g., ethnography, grounded theory) or data collection methods (e.g., interview, focus group) is recommended | Yes | |
| **Abstract** - Summary of key elements of the study using the abstract format of the intended publication; typically includes background, purpose, methods, results, and conclusions | Yes | |
| **Problem formulation -** Description and significance | Yes | |

| | |
|---|---|
| of the problem/phenomenon studied; review of relevant theory and empirical work; problem statement | |
| **Purpose or research questio**n - Purpose of the study and specific objectives or questions | Yes |
| **Qualitative approach and research paradigm** - Qualitative approach (e.g., ethnography, grounded theory, case study, phenomenology, narrative research) and guiding theory if appropriate; identifying the research paradigm (e.g., postpositivist, constructivist/ interpretivist) is also recommended; rationale** | No |
| **Researcher characteristics and reflexivity** - Researchers' characteristics that may influence the research, including personal attributes, qualifications/experience, relationship with participants, assumptions, and/or presuppositions; potential or actual interaction between researchers' characteristics and the research questions, approach, methods, results, and/or transferability | No |
| **Context** - Setting/site and salient contextual factors; rationale** | Yes |
| **Sampling strategy** - How and why research participants, documents, or events were selected; criteria for deciding when no further sampling was necessary (e.g., sampling saturation); rationale** | No |
| **Ethical issues pertaining to human subjects** - Documentation of approval by an appropriate ethics review board and participant consent, or explanation for lack thereof; other confidentiality and data security issues | No |
| **Data collection methods** - Types of data collected; details of data collection procedures including (as appropriate) start and stop dates of data collection and analysis, iterative process, triangulation of sources/methods, and modification of procedures in response to evolving study findings; rationale** | Yes |
| **Data collection instruments and technologies** - Description of instruments (e.g., interview guides, questionnaires) and devices (e.g., audio recorders) used for data collection; if/how the instrument(s) changed over the course of the study | Yes |
| **Units of study** - Number and relevant characteristics of participants, documents, or events included in the study; level of participation (could be reported in | Yes |

| | | |
|---|---|---|
| results) | | |
| **Data processing -** Methods for processing data prior to and during analysis, including transcription, data entry, data management and security, verification of data integrity, data coding, and anonymization/de-identification of excerpts | No | |
| **Data analysis -** Process by which inferences, themes, etc., were identified and developed, including the researchers involved in data analysis; usually references a specific paradigm or approach; rationale** | Yes | |
| **Techniques to enhance trustworthiness -** Techniques to enhance trustworthiness and credibility of data analysis (e.g., member checking, audit trail, triangulation); rationale** | No | |
| **Synthesis and interpretation -** Main findings (e.g., interpretations, inferences, and themes); might include development of a theory or model, or integration with prior research or theory | Yes | |
| **Links to empirical data -** Evidence (e.g., quotes, field notes, text excerpts, photographs) to substantiate analytic findings | Yes | |
| **Integration with prior work, implications, transferability, and contribution(s) to the field -** Short summary of main findings; explanation of how findings and conclusions connect to, support, elaborate on, or challenge conclusions of earlier scholarship; discussion of scope of application/generalizability; identification of unique contribution(s) to scholarship in a discipline or field | Yes | |
| **Limitations -** Trustworthiness and limitations of findings | Yes | |
| **Conflicts of interest -** Potential sources of influence or perceived influence on study conduct and conclusions; how these were managed | No | |
| **Funding -** Sources of funding and other support; role of funders in data collection, interpretation, and reporting | No | 13/21 |

| **Article Name and Author: Study 7** | **Briefing-debriefing: using a reflexive organizational learning model from the military to enhance the performance of surgical teams** | **Vashdi et al. (2007)** |
|---|---|---|
| **Title -** Concise description of the nature and topic of the study Identifying the study as qualitative or indicating the approach (e.g., ethnography, | Yes | |

| | |
|---|---|
| grounded theory) or data collection methods (e.g., interview, focus group) is recommended | |
| **Abstract** - Summary of key elements of the study using the abstract format of the intended publication; typically includes background, purpose, methods, results, and conclusions | Yes |
| **Problem formulation** - Description and significance of the problem/phenomenon studied; review of relevant theory and empirical work; problem statement | Yes |
| **Purpose or research questio**n - Purpose of the study and specific objectives or questions | Yes |
| **Qualitative approach and research paradigm** - Qualitative approach (e.g., ethnography, grounded theory, case study, phenomenology, narrative research) and guiding theory if appropriate; identifying the research paradigm (e.g., postpositivist, constructivist/ interpretivist) is also recommended; rationale** | Yes |
| **Researcher characteristics and reflexivity** - Researchers' characteristics that may influence the research, including personal attributes, qualifications/experience, relationship with participants, assumptions, and/or presuppositions; potential or actual interaction between researchers' characteristics and the research questions, approach, methods, results, and/or transferability | No |
| **Context** - Setting/site and salient contextual factors; rationale** | Yes |
| **Sampling strategy** - How and why research participants, documents, or events were selected; criteria for deciding when no further sampling was necessary (e.g., sampling saturation); rationale** | Yes |
| **Ethical issues pertaining to human subjects** - Documentation of approval by an appropriate ethics review board and participant consent, or explanation for lack thereof; other confidentiality and data security issues | No |
| **Data collection methods** - Types of data collected; details of data collection procedures including (as appropriate) start and stop dates of data collection and analysis, iterative process, triangulation of sources/methods, and modification of procedures in response to evolving study findings; rationale** | Yes |
| **Data collection instruments and technologies** - | Yes |

| | | |
|---|---|---|
| Description of instruments (e.g., interview guides, questionnaires) and devices (e.g., audio recorders) used for data collection; if/how the instrument(s) changed over the course of the study | | |
| **Units of study -** Number and relevant characteristics of participants, documents, or events included in the study; level of participation (could be reported in results) | No | |
| **Data processing -** Methods for processing data prior to and during analysis, including transcription, data entry, data management and security, verification of data integrity, data coding, and anonymization/de-identification of excerpts | Yes | |
| **Data analysis -** Process by which inferences, themes, etc., were identified and developed, including the researchers involved in data analysis; usually references a specific paradigm or approach; rationale** | Yes | |
| **Techniques to enhance trustworthiness -** Techniques to enhance trustworthiness and credibility of data analysis (e.g., member checking, audit trail, triangulation); rationale** | No | |
| **Synthesis and interpretation -** Main findings (e.g., interpretations, inferences, and themes); might include development of a theory or model, or integration with prior research or theory | Yes | |
| **Links to empirical data -** Evidence (e.g., quotes, field notes, text excerpts, photographs) to substantiate analytic findings | Yes | |
| **Integration with prior work, implications, transferability, and contribution(s) to the field -** Short summary of main findings; explanation of how findings and conclusions connect to, support, elaborate on, or challenge conclusions of earlier scholarship; discussion of scope of application/generalizability; identification of unique contribution(s) to scholarship in a discipline or field | Yes | |
| **Limitations -** Trustworthiness and limitations of findings | Yes | |
| **Conflicts of interest -** Potential sources of influence or perceived influence on study conduct and conclusions; how these were managed | No | |
| **Funding -** Sources of funding and other support; role of funders in data collection, interpretation, and reporting | No | 15/21 |

| **Article Name and Author: Study 8** | **Surfacing safety hazards using stanardized operating room briefings and debriefings at a large regional medical center** | **Bandari et al. (2012)** |
|---|---|---|
| **Title -** Concise description of the nature and topic of the study Identifying the study as qualitative or indicating the approach (e.g., ethnography, grounded theory) or data collection methods (e.g., interview, focus group) is recommended | Yes | |
| **Abstract** - Summary of key elements of the study using the abstract format of the intended publication; typically includes background, purpose, methods, results, and conclusions | Yes | |
| **Problem formulation -** Description and significance of the problem/phenomenon studied; review of relevant theory and empirical work; problem statement | Yes | |
| **Purpose or research questio**n **-** Purpose of the study and specific objectives or questions | Yes | |
| **Qualitative approach and research paradigm -** Qualitative approach (e.g., ethnography, grounded theory, case study, phenomenology, narrative research) and guiding theory if appropriate; identifying the research paradigm (e.g., postpositivist, constructivist/ interpretivist) is also recommended; rationale** | No | |
| **Researcher characteristics and reflexivity -** Researchers' characteristics that may influence the research, including personal attributes, qualifications/experience, relationship with participants, assumptions, and/or presuppositions; potential or actual interaction between researchers' characteristics and the research questions, approach, methods, results, and/or transferability | No | |
| **Context -** Setting/site and salient contextual factors; rationale** | Yes | |
| **Sampling strategy -** How and why research participants, documents, or events were selected; criteria for deciding when no further sampling was necessary (e.g., sampling saturation); rationale** | Yes | |
| **Ethical issues pertaining to human subjects -** Documentation of approval by an appropriate ethics review board and participant consent, or explanation for lack thereof; other confidentiality and data security issues | No | |

| | |
|---|---|
| **Data collection methods -** Types of data collected; details of data collection procedures including (as appropriate) start and stop dates of data collection and analysis, iterative process, triangulation of sources/methods, and modification of procedures in response to evolving study findings; rationale** | Yes |
| **Data collection instruments and technologies -** Description of instruments (e.g., interview guides, questionnaires) and devices (e.g., audio recorders) used for data collection; if/how the instrument(s) changed over the course of the study | Yes |
| **Units of study -** Number and relevant characteristics of participants, documents, or events included in the study; level of participation (could be reported in results) | Yes |
| **Data processing -** Methods for processing data prior to and during analysis, including transcription, data entry, data management and security, verification of data integrity, data coding, and anonymization/de-identification of excerpts | Yes |
| **Data analysis -** Process by which inferences, themes, etc., were identified and developed, including the researchers involved in data analysis; usually references a specific paradigm or approach; rationale** | No |
| **Techniques to enhance trustworthiness -** Techniques to enhance trustworthiness and credibility of data analysis (e.g., member checking, audit trail, triangulation); rationale** | No |
| **Synthesis and interpretation -** Main findings (e.g., interpretations, inferences, and themes); might include development of a theory or model, or integration with prior research or theory | Yes |
| **Links to empirical data -** Evidence (e.g., quotes, field notes, text excerpts, photographs) to substantiate analytic findings | Yes |
| **Integration with prior work, implications, transferability, and contribution(s) to the field -** Short summary of main findings; explanation of how findings and conclusions connect to, support, elaborate on, or challenge conclusions of earlier scholarship; discussion of scope of application/generalizability; identification of unique contribution(s) to scholarship in a discipline or field | Yes |
| **Limitations -** Trustworthiness and limitations of | Yes |

| | | |
|---|---|---|
| findings | | |
| **Conflicts of interest -** Potential sources of influence or perceived influence on study conduct and conclusions; how these were managed | No | |
| **Funding -** Sources of funding and other support; role of funders in data collection, interpretation, and reporting | No | 14/21 |
| **Article Name and Author: Study 9** | **Implementing standardized operating room briefings and debriefings at a large regional medical center** | **Berenholtz et al. (2009)** |
| **Abstract** - Summary of key elements of the study using the abstract format of the intended publication; typically includes background, purpose, methods, results, and conclusions | Yes | |
| **Problem formulation -** Description and significance of the problem/phenomenon studied; review of relevant theory and empirical work; problem statement | Yes | |
| **Purpose or research questio**n **-** Purpose of the study and specific objectives or questions | Yes | |
| **Qualitative approach and research paradigm -** Qualitative approach (e.g., ethnography, grounded theory, case study, phenomenology, narrative research) and guiding theory if appropriate; identifying the research paradigm (e.g., postpositivist, constructivist/ interpretivist) is also recommended; rationale** | Yes | |
| **Researcher characteristics and reflexivity -** Researchers' characteristics that may influence the research, including personal attributes, qualifications/experience, relationship with participants, assumptions, and/or presuppositions; potential or actual interaction between researchers' characteristics and the research questions, approach, methods, results, and/or transferability | No | |
| **Context -** Setting/site and salient contextual factors; rationale** | No | |
| **Sampling strategy -** How and why research participants, documents, or events were selected; criteria for deciding when no further sampling was necessary (e.g., sampling saturation); rationale** | Yes | |
| **Ethical issues pertaining to human subjects -** Documentation of approval by an appropriate ethics review board and participant consent, or explanation for lack thereof; other confidentiality and data | Yes | |

| | |
|---|---|
| security issues | |
| **Data collection methods -** Types of data collected; details of data collection procedures including (as appropriate) start and stop dates of data collection and analysis, iterative process, triangulation of sources/methods, and modification of procedures in response to evolving study findings; rationale** | Yes |
| **Data collection instruments and technologies -** Description of instruments (e.g., interview guides, questionnaires) and devices (e.g., audio recorders) used for data collection; if/how the instrument(s) changed over the course of the study | Yes |
| **Units of study -** Number and relevant characteristics of participants, documents, or events included in the study; level of participation (could be reported in results) | Yes |
| **Data processing -** Methods for processing data prior to and during analysis, including transcription, data entry, data management and security, verification of data integrity, data coding, and anonymization/de-identification of excerpts | Yes |
| **Data analysis -** Process by which inferences, themes, etc., were identified and developed, including the researchers involved in data analysis; usually references a specific paradigm or approach; rationale** | No |
| **Techniques to enhance trustworthiness -** Techniques to enhance trustworthiness and credibility of data analysis (e.g., member checking, audit trail, triangulation); rationale** | No |
| **Synthesis and interpretation -** Main findings (e.g., interpretations, inferences, and themes); might include development of a theory or model, or integration with prior research or theory | Yes |
| **Links to empirical data -** Evidence (e.g., quotes, field notes, text excerpts, photographs) to substantiate analytic findings | No |
| **Integration with prior work, implications, transferability, and contribution(s) to the field -** Short summary of main findings; explanation of how findings and conclusions connect to, support, elaborate on, or challenge conclusions of earlier scholarship; discussion of scope of application/generalizability; identification of unique contribution(s) to scholarship in a discipline or field | Yes |

| | | |
|---|---|---|
| **Limitations -** Trustworthiness and limitations of findings | Yes | |
| **Conflicts of interest -** Potential sources of influence or perceived influence on study conduct and conclusions; how these were managed | No | |
| **Funding -** Sources of funding and other support; role of funders in data collection, interpretation, and reporting | No | 14/21 |
| **Article Name and Author: Study 10** | **Use of briefings and debriefings as a tool in improving teamwork, efficiency, and communication in the operating theatre** | **Bethune et al. (2011)** |
| **Title -** Concise description of the nature and topic of the study Identifying the study as qualitative or indicating the approach (e.g., ethnography, grounded theory) or data collection methods (e.g., interview, focus group) is recommended | No | |
| **Abstract** - Summary of key elements of the study using the abstract format of the intended publication; typically includes background, purpose, methods, results, and conclusions | Yes | |
| **Problem formulation -** Description and significance of the problem/phenomenon studied; review of relevant theory and empirical work; problem statement | Yes | |
| **Purpose or research questio**n **-** Purpose of the study and specific objectives or questions | Yes | |
| **Qualitative approach and research paradigm -** Qualitative approach (e.g., ethnography, grounded theory, case study, phenomenology, narrative research) and guiding theory if appropriate; identifying the research paradigm (e.g., postpositivist, constructivist/ interpretivist) is also recommended; rationale** | Yes | |
| **Researcher characteristics and reflexivity -** Researchers' characteristics that may influence the research, including personal attributes, qualifications/experience, relationship with participants, assumptions, and/or presuppositions; potential or actual interaction between researchers' characteristics and the research questions, approach, methods, results, and/or transferability | No | |
| **Context -** Setting/site and salient contextual factors; rationale** | Yes | |
| **Sampling strategy -** How and why research | No | |

| | |
|---|---|
| participants, documents, or events were selected; criteria for deciding when no further sampling was necessary (e.g., sampling saturation); rationale** | |
| **Ethical issues pertaining to human subjects -** Documentation of approval by an appropriate ethics review board and participant consent, or explanation for lack thereof; other confidentiality and data security issues | No |
| **Data collection methods -** Types of data collected; details of data collection procedures including (as appropriate) start and stop dates of data collection and analysis, iterative process, triangulation of sources/methods, and modification of procedures in response to evolving study findings; rationale** | Yes |
| **Data collection instruments and technologies -** Description of instruments (e.g., interview guides, questionnaires) and devices (e.g., audio recorders) used for data collection; if/how the instrument(s) changed over the course of the study | Yes |
| **Units of study -** Number and relevant characteristics of participants, documents, or events included in the study; level of participation (could be reported in results) | Yes |
| **Data processing -** Methods for processing data prior to and during analysis, including transcription, data entry, data management and security, verification of data integrity, data coding, and anonymization/de-identification of excerpts | No |
| **Data analysis -** Process by which inferences, themes, etc., were identified and developed, including the researchers involved in data analysis; usually references a specific paradigm or approach; rationale** | No |
| **Techniques to enhance trustworthiness -** Techniques to enhance trustworthiness and credibility of data analysis (e.g., member checking, audit trail, triangulation); rationale** | No |
| **Synthesis and interpretation -** Main findings (e.g., interpretations, inferences, and themes); might include development of a theory or model, or integration with prior research or theory | Yes |
| **Links to empirical data -** Evidence (e.g., quotes, field notes, text excerpts, photographs) to substantiate analytic findings | Yes |
| **Integration with prior work, implications,** | Yes |

| | | |
|---|---|---|
| **transferability, and contribution(s) to the field -** Short summary of main findings; explanation of how findings and conclusions connect to, support, elaborate on, or challenge conclusions of earlier scholarship; discussion of scope of application/generalizability; identification of unique contribution(s) to scholarship in a discipline or field | | |
| **Limitations** - Trustworthiness and limitations of findings | No | |
| **Conflicts of interest -** Potential sources of influence or perceived influence on study conduct and conclusions; how these were managed | Yes | |
| **Funding -** Sources of funding and other support; role of funders in data collection, interpretation, and reporting | No | 13/21 |
| **Article Name and Author: Study 11** | **Implementation of surgical debriefing programs in large health systems: an exploratory qualitative analysis** | **Brindle et al. (2018)** |
| **Abstract** - Summary of key elements of the study using the abstract format of the intended publication; typically includes background, purpose, methods, results, and conclusions | Yes | |
| **Problem formulation -** Description and significance of the problem/phenomenon studied; review of relevant theory and empirical work; problem statement | Yes | |
| **Purpose or research questio**n - Purpose of the study and specific objectives or questions | Yes | |
| **Qualitative approach and research paradigm -** Qualitative approach (e.g., ethnography, grounded theory, case study, phenomenology, narrative research) and guiding theory if appropriate; identifying the research paradigm (e.g., postpositivist, constructivist/ interpretivist) is also recommended; rationale** | Yes | |
| **Researcher characteristics and reflexivity -** Researchers' characteristics that may influence the research, including personal attributes, qualifications/experience, relationship with participants, assumptions, and/or presuppositions; potential or actual interaction between researchers' characteristics and the research questions, approach, methods, results, and/or transferability | Yes | |
| **Context -** Setting/site and salient contextual factors; rationale** | No | |

| | |
|---|---|
| **Sampling strategy -** How and why research participants, documents, or events were selected; criteria for deciding when no further sampling was necessary (e.g., sampling saturation); rationale** | Yes |
| **Ethical issues pertaining to human subjects -** Documentation of approval by an appropriate ethics review board and participant consent, or explanation for lack thereof; other confidentiality and data security issues | Yes |
| **Data collection methods -** Types of data collected; details of data collection procedures including (as appropriate) start and stop dates of data collection and analysis, iterative process, triangulation of sources/methods, and modification of procedures in response to evolving study findings; rationale** | Yes |
| **Data collection instruments and technologies -** Description of instruments (e.g., interview guides, questionnaires) and devices (e.g., audio recorders) used for data collection; if/how the instrument(s) changed over the course of the study | Yes |
| **Units of study -** Number and relevant characteristics of participants, documents, or events included in the study; level of participation (could be reported in results) | Yes |
| **Data processing -** Methods for processing data prior to and during analysis, including transcription, data entry, data management and security, verification of data integrity, data coding, and anonymization/de-identification of excerpts | No |
| **Data analysis -** Process by which inferences, themes, etc., were identified and developed, including the researchers involved in data analysis; usually references a specific paradigm or approach; rationale** | Yes |
| **Techniques to enhance trustworthiness -** Techniques to enhance trustworthiness and credibility of data analysis (e.g., member checking, audit trail, triangulation); rationale** | Yes |
| **Synthesis and interpretation -** Main findings (e.g., interpretations, inferences, and themes); might include development of a theory or model, or integration with prior research or theory | No |
| **Links to empirical data -** Evidence (e.g., quotes, field notes, text excerpts, photographs) to substantiate analytic findings | Yes |

| | | |
|---|---|---|
| **Integration with prior work, implications, transferability, and contribution(s) to the field -** Short summary of main findings; explanation of how findings and conclusions connect to, support, elaborate on, or challenge conclusions of earlier scholarship; discussion of scope of application/generalizability; identification of unique contribution(s) to scholarship in a discipline or field | Yes | |
| **Limitations -** Trustworthiness and limitations of findings | No | |
| **Conflicts of interest -** Potential sources of influence or perceived influence on study conduct and conclusions; how these were managed | Yes | |
| **Funding -** Sources of funding and other support; role of funders in data collection, interpretation, and reporting | Yes | 17/21 |

| **Article Name and Author: Study 12** | **Debriefing in the OR: A quality improvement project** | **Finch et al. (2019)** |
|---|---|---|
| **Title -** Concise description of the nature and topic of the study Identifying the study as qualitative or indicating the approach (e.g., ethnography, grounded theory) or data collection methods (e.g., interview, focus group) is recommended | Yes | |
| **Abstract** - Summary of key elements of the study using the abstract format of the intended publication; typically includes background, purpose, methods, results, and conclusions | Yes | |
| **Problem formulation -** Description and significance of the problem/phenomenon studied; review of relevant theory and empirical work; problem statement | Yes | |
| **Purpose or research questio**n - Purpose of the study and specific objectives or questions | Yes | |
| **Qualitative approach and research paradigm -** Qualitative approach (e.g., ethnography, grounded theory, case study, phenomenology, narrative research) and guiding theory if appropriate; identifying the research paradigm (e.g., postpositivist, constructivist/ interpretivist) is also recommended; rationale** | No | |
| **Researcher characteristics and reflexivity -** Researchers' characteristics that may influence the research, including personal attributes, qualifications/experience, relationship with | No | |

| | |
|---|---|
| participants, assumptions, and/or presuppositions; potential or actual interaction between researchers' characteristics and the research questions, approach, methods, results, and/or transferability | |
| **Context** - Setting/site and salient contextual factors; rationale** | Yes |
| **Sampling strategy** - How and why research participants, documents, or events were selected; criteria for deciding when no further sampling was necessary (e.g., sampling saturation); rationale** | Yes |
| **Ethical issues pertaining to human subjects** - Documentation of approval by an appropriate ethics review board and participant consent, or explanation for lack thereof; other confidentiality and data security issues | Yes |
| **Data collection methods** - Types of data collected; details of data collection procedures including (as appropriate) start and stop dates of data collection and analysis, iterative process, triangulation of sources/methods, and modification of procedures in response to evolving study findings; rationale** | Yes |
| **Data collection instruments and technologies** - Description of instruments (e.g., interview guides, questionnaires) and devices (e.g., audio recorders) used for data collection; if/how the instrument(s) changed over the course of the study | Yes |
| **Units of study** - Number and relevant characteristics of participants, documents, or events included in the study; level of participation (could be reported in results) | No |
| **Data processing** - Methods for processing data prior to and during analysis, including transcription, data entry, data management and security, verification of data integrity, data coding, and anonymization/de-identification of excerpts | Yes |
| **Data analysis** - Process by which inferences, themes, etc., were identified and developed, including the researchers involved in data analysis; usually references a specific paradigm or approach; rationale** | No |
| **Techniques to enhance trustworthiness** - Techniques to enhance trustworthiness and credibility of data analysis (e.g., member checking, audit trail, triangulation); rationale** | No |
| **Synthesis and interpretation** - Main findings (e.g., | Yes |

| | | |
|---|---|---|
| interpretations, inferences, and themes); might include development of a theory or model, or integration with prior research or theory | | |
| **Links to empirical data** - Evidence (e.g., quotes, field notes, text excerpts, photographs) to substantiate analytic findings | No | |
| **Integration with prior work, implications, transferability, and contribution(s) to the field -** Short summary of main findings; explanation of how findings and conclusions connect to, support, elaborate on, or challenge conclusions of earlier scholarship; discussion of scope of application/generalizability; identification of unique contribution(s) to scholarship in a discipline or field | Yes | |
| **Limitations -** Trustworthiness and limitations of findings | No | |
| **Conflicts of interest -** Potential sources of influence or perceived influence on study conduct and conclusions; how these were managed | Yes | |
| **Funding -** Sources of funding and other support; role of funders in data collection, interpretation, and reporting | No | 13/21 |

| **Article Name and Author: Study 13** | **Coaching to improve the quality of communication during briefings and debriefings** | **Kleiner et al. (2014)** |
|---|---|---|
| **Abstract** - Summary of key elements of the study using the abstract format of the intended publication; typically includes background, purpose, methods, results, and conclusions | Yes | |
| **Problem formulation** - Description and significance of the problem/phenomenon studied; review of relevant theory and empirical work; problem statement | Yes | |
| **Purpose or research questio**n - Purpose of the study and specific objectives or questions | Yes | |
| **Qualitative approach and research paradigm** - Qualitative approach (e.g., ethnography, grounded theory, case study, phenomenology, narrative research) and guiding theory if appropriate; identifying the research paradigm (e.g., postpositivist, constructivist/ interpretivist) is also recommended; rationale** | Yes | |
| **Researcher characteristics and reflexivity** - Researchers' characteristics that may influence the research, including personal attributes, | No | |

| | |
|---|---|
| qualifications/experience, relationship with participants, assumptions, and/or presuppositions; potential or actual interaction between researchers' characteristics and the research questions, approach, methods, results, and/or transferability | |
| **Context** - Setting/site and salient contextual factors; rationale** | Yes |
| **Sampling strategy** - How and why research participants, documents, or events were selected; criteria for deciding when no further sampling was necessary (e.g., sampling saturation); rationale** | Yes |
| **Ethical issues pertaining to human subjects** - Documentation of approval by an appropriate ethics review board and participant consent, or explanation for lack thereof; other confidentiality and data security issues | Yes |
| **Data collection methods** - Types of data collected; details of data collection procedures including (as appropriate) start and stop dates of data collection and analysis, iterative process, triangulation of sources/methods, and modification of procedures in response to evolving study findings; rationale** | Yes |
| **Data collection instruments and technologies** - Description of instruments (e.g., interview guides, questionnaires) and devices (e.g., audio recorders) used for data collection; if/how the instrument(s) changed over the course of the study | Yes |
| **Units of study** - Number and relevant characteristics of participants, documents, or events included in the study; level of participation (could be reported in results) | Yes |
| **Data processing** - Methods for processing data prior to and during analysis, including transcription, data entry, data management and security, verification of data integrity, data coding, and anonymization/de-identification of excerpts | Yes |
| **Data analysis** - Process by which inferences, themes, etc., were identified and developed, including the researchers involved in data analysis; usually references a specific paradigm or approach; rationale** | No |
| **Techniques to enhance trustworthiness** - Techniques to enhance trustworthiness and credibility of data analysis (e.g., member checking, audit trail, triangulation); rationale** | Yes |

| | | |
|---|---|---|
| **Synthesis and interpretation** - Main findings (e.g., interpretations, inferences, and themes); might include development of a theory or model, or integration with prior research or theory | Yes | |
| **Links to empirical data** - Evidence (e.g., quotes, field notes, text excerpts, photographs) to substantiate analytic findings | No | |
| **Integration with prior work, implications, transferability, and contribution(s) to the field -** Short summary of main findings; explanation of how findings and conclusions connect to, support, elaborate on, or challenge conclusions of earlier scholarship; discussion of scope of application/generalizability; identification of unique contribution(s) to scholarship in a discipline or field | Yes | |
| **Limitations -** Trustworthiness and limitations of findings | Yes | |
| **Conflicts of interest -** Potential sources of influence or perceived influence on study conduct and conclusions; how these were managed | Yes | |
| **Funding -** Sources of funding and other support; role of funders in data collection, interpretation, and reporting | Yes | 18/21 |

| **Article Name and Author: Study 14** | **Effects of perioperative briefing and debriefing on patient safety: a prospective intervention study** | **Leong et al. (2017)** |
|---|---|---|
| **Title -** Concise description of the nature and topic of the study Identifying the study as qualitative or indicating the approach (e.g., ethnography, grounded theory) or data collection methods (e.g., interview, focus group) is recommended | Yes | |
| **Abstract** - Summary of key elements of the study using the abstract format of the intended publication; typically includes background, purpose, methods, results, and conclusions | Yes | |
| **Problem formulation -** Description and significance of the problem/phenomenon studied; review of relevant theory and empirical work; problem statement | Yes | |
| **Purpose or research questio**n - Purpose of the study and specific objectives or questions | Yes | |
| **Qualitative approach and research paradigm -** Qualitative approach (e.g., ethnography, grounded theory, case study, phenomenology, narrative research) and guiding theory if appropriate; | No | |

| | |
|---|---|
| identifying the research paradigm (e.g., postpositivist, constructivist/ interpretivist) is also recommended; rationale** | |
| **Researcher characteristics and reflexivity -** Researchers' characteristics that may influence the research, including personal attributes, qualifications/experience, relationship with participants, assumptions, and/or presuppositions; potential or actual interaction between researchers' characteristics and the research questions, approach, methods, results, and/or transferability | No |
| **Context -** Setting/site and salient contextual factors; rationale** | Yes |
| **Sampling strategy -** How and why research participants, documents, or events were selected; criteria for deciding when no further sampling was necessary (e.g., sampling saturation); rationale** | Yes |
| **Ethical issues pertaining to human subjects -** Documentation of approval by an appropriate ethics review board and participant consent, or explanation for lack thereof; other confidentiality and data security issues | Yes |
| **Data collection methods -** Types of data collected; details of data collection procedures including (as appropriate) start and stop dates of data collection and analysis, iterative process, triangulation of sources/methods, and modification of procedures in response to evolving study findings; rationale** | Yes |
| **Data collection instruments and technologies -** Description of instruments (e.g., interview guides, questionnaires) and devices (e.g., audio recorders) used for data collection; if/how the instrument(s) changed over the course of the study | Yes |
| **Units of study -** Number and relevant characteristics of participants, documents, or events included in the study; level of participation (could be reported in results) | Yes |
| **Data processing -** Methods for processing data prior to and during analysis, including transcription, data entry, data management and security, verification of data integrity, data coding, and anonymization/de-identification of excerpts | Yes |
| **Data analysis -** Process by which inferences, themes, etc., were identified and developed, including the researchers involved in data analysis; usually | No |

| | | |
|---|---|---|
| references a specific paradigm or approach; rationale** | | |
| **Techniques to enhance trustworthiness -** Techniques to enhance trustworthiness and credibility of data analysis (e.g., member checking, audit trail, triangulation); rationale** | No | |
| **Synthesis and interpretation -** Main findings (e.g., interpretations, inferences, and themes); might include development of a theory or model, or integration with prior research or theory | Yes | |
| **Links to empirical data -** Evidence (e.g., quotes, field notes, text excerpts, photographs) to substantiate analytic findings | No | |
| **Integration with prior work, implications, transferability, and contribution(s) to the field -** Short summary of main findings; explanation of how findings and conclusions connect to, support, elaborate on, or challenge conclusions of earlier scholarship; discussion of scope of application/generalizability; identification of unique contribution(s) to scholarship in a discipline or field | Yes | |
| **Limitations -** Trustworthiness and limitations of findings | Yes | |
| **Conflicts of interest -** Potential sources of influence or perceived influence on study conduct and conclusions; how these were managed | Yes | |
| **Funding -** Sources of funding and other support; role of funders in data collection, interpretation, and reporting | No | 15/21 |

| **Article Name and Author: Study 15** | **Changing operating room culture: implementation of a postoperative debrief and improved safety culture** | **Magill et al. (2017)** |
|---|---|---|
| **Title -** Concise description of the nature and topic of the study Identifying the study as qualitative or indicating the approach (e.g., ethnography, grounded theory) or data collection methods (e.g., interview, focus group) is recommended | Yes | |
| **Abstract -** Summary of key elements of the study using the abstract format of the intended publication; typically includes background, purpose, methods, results, and conclusions | Yes | |
| **Problem formulation -** Description and significance of the problem/phenomenon studied; review of relevant theory and empirical work; problem statement | Yes | |

| | |
|---|---|
| **Purpose or research questio**n - Purpose of the study and specific objectives or questions | Yes |
| **Qualitative approach and research paradigm -** Qualitative approach (e.g., ethnography, grounded theory, case study, phenomenology, narrative research) and guiding theory if appropriate; identifying the research paradigm (e.g., postpositivist, constructivist/ interpretivist) is also recommended; rationale** | No |
| **Researcher characteristics and reflexivity -** Researchers' characteristics that may influence the research, including personal attributes, qualifications/experience, relationship with participants, assumptions, and/or presuppositions; potential or actual interaction between researchers' characteristics and the research questions, approach, methods, results, and/or transferability | No |
| **Context -** Setting/site and salient contextual factors; rationale** | Yes |
| **Sampling strategy -** How and why research participants, documents, or events were selected; criteria for deciding when no further sampling was necessary (e.g., sampling saturation); rationale** | Yes |
| **Ethical issues pertaining to human subjects -** Documentation of approval by an appropriate ethics review board and participant consent, or explanation for lack thereof; other confidentiality and data security issues | No |
| **Data collection methods -** Types of data collected; details of data collection procedures including (as appropriate) start and stop dates of data collection and analysis, iterative process, triangulation of sources/methods, and modification of procedures in response to evolving study findings; rationale** | Yes |
| **Data collection instruments and technologies -** Description of instruments (e.g., interview guides, questionnaires) and devices (e.g., audio recorders) used for data collection; if/how the instrument(s) changed over the course of the study | Yes |
| **Units of study -** Number and relevant characteristics of participants, documents, or events included in the study; level of participation (could be reported in results) | Yes |
| **Data processing -** Methods for processing data prior to and during analysis, including transcription, data | Yes |

| | | |
|---|---|---|
| entry, data management and security, verification of data integrity, data coding, and anonymization/de-identification of excerpts | | |
| **Data analysis** - Process by which inferences, themes, etc., were identified and developed, including the researchers involved in data analysis; usually references a specific paradigm or approach; rationale** | No | |
| **Techniques to enhance trustworthiness** - Techniques to enhance trustworthiness and credibility of data analysis (e.g., member checking, audit trail, triangulation); rationale** | No | |
| **Synthesis and interpretation** - Main findings (e.g., interpretations, inferences, and themes); might include development of a theory or model, or integration with prior research or theory | Yes | |
| **Links to empirical data** - Evidence (e.g., quotes, field notes, text excerpts, photographs) to substantiate analytic findings | Yes | |
| **Integration with prior work, implications, transferability, and contribution(s) to the field -** Short summary of main findings; explanation of how findings and conclusions connect to, support, elaborate on, or challenge conclusions of earlier scholarship; discussion of scope of application/generalizability; identification of unique contribution(s) to scholarship in a discipline or field | Yes | |
| **Limitations** - Trustworthiness and limitations of findings | Yes | |
| **Conflicts of interest** - Potential sources of influence or perceived influence on study conduct and conclusions; how these were managed | Yes | |
| **Funding** - Sources of funding and other support; role of funders in data collection, interpretation, and reporting | No | 15/21 |

| **Article Name and Author: Study 16** | **Okay, let's talk - short debriefings in the operating room** | **Mundt et al. (2020)** |
|---|---|---|
| **Title** - Concise description of the nature and topic of the study Identifying the study as qualitative or indicating the approach (e.g., ethnography, grounded theory) or data collection methods (e.g., interview, focus group) is recommended | Yes | |
| **Abstract** - Summary of key elements of the study using the abstract format of the intended | Yes | |

| | |
|---|---|
| publication; typically includes background, purpose, methods, results, and conclusions | |
| **Problem formulation** - Description and significance of the problem/phenomenon studied; review of relevant theory and empirical work; problem statement | Yes |
| **Purpose or research questio**n - Purpose of the study and specific objectives or questions | Yes |
| **Qualitative approach and research paradigm** - Qualitative approach (e.g., ethnography, grounded theory, case study, phenomenology, narrative research) and guiding theory if appropriate; identifying the research paradigm (e.g., postpositivist, constructivist/ interpretivist) is also recommended; rationale** | Yes |
| **Researcher characteristics and reflexivity** - Researchers' characteristics that may influence the research, including personal attributes, qualifications/experience, relationship with participants, assumptions, and/or presuppositions; potential or actual interaction between researchers' characteristics and the research questions, approach, methods, results, and/or transferability | No |
| **Context** - Setting/site and salient contextual factors; rationale** | Yes |
| **Sampling strategy** - How and why research participants, documents, or events were selected; criteria for deciding when no further sampling was necessary (e.g., sampling saturation); rationale** | Yes |
| **Ethical issues pertaining to human subjects** - Documentation of approval by an appropriate ethics review board and participant consent, or explanation for lack thereof; other confidentiality and data security issues | Yes |
| **Data collection methods** - Types of data collected; details of data collection procedures including (as appropriate) start and stop dates of data collection and analysis, iterative process, triangulation of sources/methods, and modification of procedures in response to evolving study findings; rationale** | Yes |
| **Data collection instruments and technologies** - Description of instruments (e.g., interview guides, questionnaires) and devices (e.g., audio recorders) used for data collection; if/how the instrument(s) changed over the course of the study | Yes |

| | | |
|---|---|---|
| **Units of study** - Number and relevant characteristics of participants, documents, or events included in the study; level of participation (could be reported in results) | Yes | |
| **Data processing** - Methods for processing data prior to and during analysis, including transcription, data entry, data management and security, verification of data integrity, data coding, and anonymization/de-identification of excerpts | Yes | |
| **Data analysis** - Process by which inferences, themes, etc., were identified and developed, including the researchers involved in data analysis; usually references a specific paradigm or approach; rationale** | Yes | |
| **Techniques to enhance trustworthiness** - Techniques to enhance trustworthiness and credibility of data analysis (e.g., member checking, audit trail, triangulation); rationale** | No | |
| **Synthesis and interpretation** - Main findings (e.g., interpretations, inferences, and themes); might include development of a theory or model, or integration with prior research or theory | Yes | |
| **Links to empirical data** - Evidence (e.g., quotes, field notes, text excerpts, photographs) to substantiate analytic findings | Yes | |
| **Integration with prior work, implications, transferability, and contribution(s) to the field -** Short summary of main findings; explanation of how findings and conclusions connect to, support, elaborate on, or challenge conclusions of earlier scholarship; discussion of scope of application/generalizability; identification of unique contribution(s) to scholarship in a discipline or field | Yes | |
| **Limitations** - Trustworthiness and limitations of findings | Yes | |
| **Conflicts of interest** - Potential sources of influence or perceived influence on study conduct and conclusions; how these were managed | Yes | |
| **Funding** - Sources of funding and other support; role of funders in data collection, interpretation, and reporting | Yes | 19/21 |
| **Article Name and Author: Study 17** | **Predictors of successful implementation of preoperative briefings and postoperative debriefings after medical team** | **Paull et al. (2009)** |

| | training |
|---|---|
| **Title -** Concise description of the nature and topic of the study Identifying the study as qualitative or indicating the approach (e.g., ethnography, grounded theory) or data collection methods (e.g., interview, focus group) is recommended | Yes |
| **Abstract -** Summary of key elements of the study using the abstract format of the intended publication; typically includes background, purpose, methods, results, and conclusions | Yes |
| **Problem formulation -** Description and significance of the problem/phenomenon studied; review of relevant theory and empirical work; problem statement | Yes |
| **Purpose or research questio**n **-** Purpose of the study and specific objectives or questions | Yes |
| **Qualitative approach and research paradigm -** Qualitative approach (e.g., ethnography, grounded theory, case study, phenomenology, narrative research) and guiding theory if appropriate; identifying the research paradigm (e.g., postpositivist, constructivist/ interpretivist) is also recommended; rationale** | No |
| **Researcher characteristics and reflexivity -** Researchers' characteristics that may influence the research, including personal attributes, qualifications/experience, relationship with participants, assumptions, and/or presuppositions; potential or actual interaction between researchers' characteristics and the research questions, approach, methods, results, and/or transferability | No |
| **Context -** Setting/site and salient contextual factors; rationale** | Yes |
| **Sampling strategy -** How and why research participants, documents, or events were selected; criteria for deciding when no further sampling was necessary (e.g., sampling saturation); rationale** | Yes |
| **Ethical issues pertaining to human subjects -** Documentation of approval by an appropriate ethics review board and participant consent, or explanation for lack thereof; other confidentiality and data security issues | No |
| **Data collection methods -** Types of data collected; details of data collection procedures including (as appropriate) start and stop dates of data collection | Yes |

| | |
|---|---|
| and analysis, iterative process, triangulation of sources/methods, and modification of procedures in response to evolving study findings; rationale** | |
| **Data collection instruments and technologies -** Description of instruments (e.g., interview guides, questionnaires) and devices (e.g., audio recorders) used for data collection; if/how the instrument(s) changed over the course of the study | Yes |
| **Units of study -** Number and relevant characteristics of participants, documents, or events included in the study; level of participation (could be reported in results) | Yes |
| **Data processing -** Methods for processing data prior to and during analysis, including transcription, data entry, data management and security, verification of data integrity, data coding, and anonymization/de-identification of excerpts | Yes |
| **Data analysis -** Process by which inferences, themes, etc., were identified and developed, including the researchers involved in data analysis; usually references a specific paradigm or approach; rationale** | No |
| **Techniques to enhance trustworthiness -** Techniques to enhance trustworthiness and credibility of data analysis (e.g., member checking, audit trail, triangulation); rationale** | No |
| **Synthesis and interpretation -** Main findings (e.g., interpretations, inferences, and themes); might include development of a theory or model, or integration with prior research or theory | Yes |
| **Links to empirical data -** Evidence (e.g., quotes, field notes, text excerpts, photographs) to substantiate analytic findings | No |
| **Integration with prior work, implications, transferability, and contribution(s) to the field -** Short summary of main findings; explanation of how findings and conclusions connect to, support, elaborate on, or challenge conclusions of earlier scholarship; discussion of scope of application/generalizability; identification of unique contribution(s) to scholarship in a discipline or field | Yes |
| **Limitations -** Trustworthiness and limitations of findings | No |
| **Conflicts of interest -** Potential sources of influence or perceived influence on study conduct and | No |

| | | |
|---|---|---|
| conclusions; how these were managed | | |
| **Funding -** Sources of funding and other support; role of funders in data collection, interpretation, and reporting | No | 12/21 |
| **Article Name and Author: Study 18** | **Use of a surgical debriefing checklist to achieve higher value health care** | **Rose et al. (2018)** |
| **Title -** Concise description of the nature and topic of the study Identifying the study as qualitative or indicating the approach (e.g., ethnography, grounded theory) or data collection methods (e.g., interview, focus group) is recommended | Yes | |
| **Abstract** - Summary of key elements of the study using the abstract format of the intended publication; typically includes background, purpose, methods, results, and conclusions | Yes | |
| **Problem formulation -** Description and significance of the problem/phenomenon studied; review of relevant theory and empirical work; problem statement | Yes | |
| **Purpose or research questio**n - Purpose of the study and specific objectives or questions | Yes | |
| **Qualitative approach and research paradigm -** Qualitative approach (e.g., ethnography, grounded theory, case study, phenomenology, narrative research) and guiding theory if appropriate; identifying the research paradigm (e.g., postpositivist, constructivist/ interpretivist) is also recommended; rationale** | No | |
| **Researcher characteristics and reflexivity -** Researchers' characteristics that may influence the research, including personal attributes, qualifications/experience, relationship with participants, assumptions, and/or presuppositions; potential or actual interaction between researchers' characteristics and the research questions, approach, methods, results, and/or transferability | No | |
| **Context -** Setting/site and salient contextual factors; rationale** | Yes | |
| **Sampling strategy -** How and why research participants, documents, or events were selected; criteria for deciding when no further sampling was necessary (e.g., sampling saturation); rationale** | Yes | |
| **Ethical issues pertaining to human subjects -** Documentation of approval by an appropriate ethics | No | |

| | |
|---|---|
| review board and participant consent, or explanation for lack thereof; other confidentiality and data security issues | |
| **Data collection methods -** Types of data collected; details of data collection procedures including (as appropriate) start and stop dates of data collection and analysis, iterative process, triangulation of sources/methods, and modification of procedures in response to evolving study findings; rationale** | Yes |
| **Data collection instruments and technologies -** Description of instruments (e.g., interview guides, questionnaires) and devices (e.g., audio recorders) used for data collection; if/how the instrument(s) changed over the course of the study | Yes |
| **Units of study -** Number and relevant characteristics of participants, documents, or events included in the study; level of participation (could be reported in results) | Yes |
| **Data processing -** Methods for processing data prior to and during analysis, including transcription, data entry, data management and security, verification of data integrity, data coding, and anonymization/de-identification of excerpts | Yes |
| **Data analysis -** Process by which inferences, themes, etc., were identified and developed, including the researchers involved in data analysis; usually references a specific paradigm or approach; rationale** | No |
| **Techniques to enhance trustworthiness -** Techniques to enhance trustworthiness and credibility of data analysis (e.g., member checking, audit trail, triangulation); rationale** | No |
| **Synthesis and interpretation -** Main findings (e.g., interpretations, inferences, and themes); might include development of a theory or model, or integration with prior research or theory | Yes |
| **Links to empirical data -** Evidence (e.g., quotes, field notes, text excerpts, photographs) to substantiate analytic findings | No |
| **Integration with prior work, implications, transferability, and contribution(s) to the field -** Short summary of main findings; explanation of how findings and conclusions connect to, support, elaborate on, or challenge conclusions of earlier scholarship; discussion of scope of | Yes |

| | | |
|---|---|---|
| application/generalizability; identification of unique contribution(s) to scholarship in a discipline or field | | |
| **Limitations** - Trustworthiness and limitations of findings | Yes | |
| **Conflicts of interest** - Potential sources of influence or perceived influence on study conduct and conclusions; how these were managed | Yes | |
| **Funding** - Sources of funding and other support; role of funders in data collection, interpretation, and reporting | Yes | 15/21 |
| **Article Name and Author: Study 19** | **Long‐term effects of perioperative briefing and debriefing on team climate: A mixed‐method evaluation study** | **Schaap et al. (2020)** |
| **Title** - Concise description of the nature and topic of the study Identifying the study as qualitative or indicating the approach (e.g., ethnography, grounded theory) or data collection methods (e.g., interview, focus group) is recommended | Yes | |
| **Abstract** - Summary of key elements of the study using the abstract format of the intended publication; typically includes background, purpose, methods, results, and conclusions | Yes | |
| **Problem formulation** - Description and significance of the problem/phenomenon studied; review of relevant theory and empirical work; problem statement | Yes | |
| **Purpose or research questio**n - Purpose of the study and specific objectives or questions | Yes | |
| **Qualitative approach and research paradigm** - Qualitative approach (e.g., ethnography, grounded theory, case study, phenomenology, narrative research) and guiding theory if appropriate; identifying the research paradigm (e.g., postpositivist, constructivist/ interpretivist) is also recommended; rationale** | No | |
| **Researcher characteristics and reflexivity** - Researchers' characteristics that may influence the research, including personal attributes, qualifications/experience, relationship with participants, assumptions, and/or presuppositions; potential or actual interaction between researchers' characteristics and the research questions, approach, methods, results, and/or transferability | No | |
| **Context** - Setting/site and salient contextual factors; | Yes | |

| | |
|---|---|
| rationale** | |
| **Sampling strategy -** How and why research participants, documents, or events were selected; criteria for deciding when no further sampling was necessary (e.g., sampling saturation); rationale** | Yes |
| **Ethical issues pertaining to human subjects -** Documentation of approval by an appropriate ethics review board and participant consent, or explanation for lack thereof; other confidentiality and data security issues | Yes |
| **Data collection methods -** Types of data collected; details of data collection procedures including (as appropriate) start and stop dates of data collection and analysis, iterative process, triangulation of sources/methods, and modification of procedures in response to evolving study findings; rationale** | Yes |
| **Data collection instruments and technologies -** Description of instruments (e.g., interview guides, questionnaires) and devices (e.g., audio recorders) used for data collection; if/how the instrument(s) changed over the course of the study | Yes |
| **Units of study -** Number and relevant characteristics of participants, documents, or events included in the study; level of participation (could be reported in results) | Yes |
| **Data processing -** Methods for processing data prior to and during analysis, including transcription, data entry, data management and security, verification of data integrity, data coding, and anonymization/de-identification of excerpts | Yes |
| **Data analysis -** Process by which inferences, themes, etc., were identified and developed, including the researchers involved in data analysis; usually references a specific paradigm or approach; rationale** | Yes |
| **Techniques to enhance trustworthiness -** Techniques to enhance trustworthiness and credibility of data analysis (e.g., member checking, audit trail, triangulation); rationale** | No |
| **Synthesis and interpretation -** Main findings (e.g., interpretations, inferences, and themes); might include development of a theory or model, or integration with prior research or theory | Yes |
| **Links to empirical data -** Evidence (e.g., quotes, field notes, text excerpts, photographs) to substantiate | Yes |

| | | |
|---|---|---|
| analytic findings | | |
| **Integration with prior work, implications, transferability, and contribution(s) to the field -** Short summary of main findings; explanation of how findings and conclusions connect to, support, elaborate on, or challenge conclusions of earlier scholarship; discussion of scope of application/generalizability; identification of unique contribution(s) to scholarship in a discipline or field | Yes | |
| **Limitations -** Trustworthiness and limitations of findings | Yes | |
| **Conflicts of interest -** Potential sources of influence or perceived influence on study conduct and conclusions; how these were managed | Yes | |
| **Funding -** Sources of funding and other support; role of funders in data collection, interpretation, and reporting | No | 17/21 |
